# Effect of home blood pressure monitoring on major adverse cardiovascular events and all-cause mortality – results from the VA HOMESTEAD study

**DOI:** 10.64898/2026.10.27.26366626

**Authors:** B. C. Birkelo, K. Velusamy, N.K. Rai, B. C. Taylor, M. Abdalla, K. L. Margolis, D. Shimbo, P. Muntner, D. Nelson, A. Kaplan, A. Cutting, P.E. Drawz, A. Ishani

## Abstract

**Importance:** Home blood pressure monitoring (HBPM) has been shown to lower blood pressure (BP) in randomized trials, however its real-world effect on clinical outcomes is not known.

**Objective:** To determine the association between HBPM and clinical outcomes including cardiovascular events and mortality.

**Design:** Target trial emulation employing matched cohorts with index-date follow-up.

**Setting:** Veterans Health Administration (VHA) telehealth program.

**Participants:** Eligible Veterans had uncontrolled BP (defined as a systolic BP >140 mmHg) at a primary care visit within VHA, with a systolic BP >140 mmHg at the prior primary care visit or average systolic BP >140 mmHg over the year prior. The HBPM (intervention) group included eligible Veterans who were enrolled in a HBPM program for hypertension between January 1, 2013 and December 31, 2019. Individuals in the control group (usual care) were selected from eligible Veterans who were not enrolled in HBPM. Controls were matched with those in the intervention, in part, on demographics, baseline blood pressure and antihypertensive medication use, baseline frailty, mortality risk, and selected comorbidities.

**Exposure:** HBPM in the VHA telehealth program

**Main Outcomes:** All-cause mortality and major adverse cardiovascular events (MACE), defined as the first occurrence during follow-up of acute myocardial infarction, stroke, acute heart failure, or unstable angina.

**Results:** Among 30,203 Veterans enrolled in HBPM and 30,096 non-enrolled controls, with a median duration of 202 days of HBPM and 6.4 years of follow-up, HBPM was associated with 8.3% lower risk of mortality (hazard ratio 0.92; 95% confidence interval 0.89, 0.95) and a 4.1% higher risk of MACE (HR 1.04; 95% CI 1.01, 1.08). In analyses calibrated for negative control outcomes, there was no longer a statistically significant association between HBPM and MACE.

**Conclusions and Relevance:** HBPM as utilized in routine clinical care among a large national cohort of adults with uncontrolled hypertension was associated with reduced risk of mortality. The association between HBPM with increased risk of MACE may have been due to ascertainment differences between those enrolled and not enrolled in HBPM.

## Introduction

Hypertension is highly prevalent worldwide^1^ and a leading risk factor for mortality and cardiovascular events such as myocardial infarction, stroke, and heart failure.^2–5^ Reducing blood pressure (BP) in individuals with hypertension lowers their risks of these outcomes.^6–8^ One recommended, but underutilized, tool for achieving BP control in adults with hypertension is out-of-office BP monitoring.^9^ The 2025 American Heart Association/American College of Cardiology BP management guideline recommends out-of-office BP measurement to guide therapy for hypertension,^10^ a recommendation supported by high-quality observational data demonstrating associations between high out-of-office blood pressure and increased risks of cardiovascular events and mortality, independent of BP measured in the office setting.^11^ Home BP monitoring (HBPM) is one method of out-of-office BP monitoring and involves individuals measuring their own BP at home while resting, similar to how BP is measured in the office setting. Randomized trials show HBPM lowers in-office BP, but no large study has evaluated hard outcomes in routine care.^12–14^

The Veterans Health Administration (VHA) telehealth program has HBPM data from over 260,000 individuals with hypertension. We have previously reported on the utilization of HBPM in this diverse nationwide sample of adults with uncontrolled hypertension and found that HBPM is utilized in a small percentage of patients.^15^ In this study, we evaluated the effect of HBPM on mortality and cardiovascular events in routine care in the VHA. We hypothesized that enrollment in HBPM would lower the risk for major adverse cardiac events and all-cause mortality. We conducted a target trial emulation study in which we compared the risks of major adverse cardiac events and mortality in individuals enrolled in HBPM versus a cohort of matched controls.

## Methods

### Study Setting and Data Sources

The study was approved by the University of Minnesota Institutional Review Board (STUDY00014620) and the Minneapolis VHA Institutional Review Board (4739-B). The study followed Strengthening the Reporting of Observational Studies in Epidemiology guideline.

Data used for this study included electronic health record data (EHR) from the VHA and HBPM data. EHR data were accessed through the VA Informatics and Computing Infrastructure (VINCI) secure workspace. HBPM data were obtained from the external vendors Medtronic and Cognosante. These data included over 86 million BP measurements from 262,999 Veterans measured from 2013 through 2023, including BP measurements for those enrolled in telehealth for hypertension and other non-hypertension conditions.

### Eligibility criteria

Inclusion criteria included being a Veteran, aged 18 to 90 years, having established primary care within VHA, and a systolic BP >140 mmHg obtained at a primary care visit (index visit) with either a systolic BP >140 mmHg at the prior primary care visit or an average systolic BP >140 mmHg over the previous year. We included eligible primary care visits occurring between January 1, 2013 and December 31, 2019. Exclusion criteria included receipt of any home telehealth prior to the first eligible primary care visit. We also excluded primary care visits with providers that did not match the Veteran’s primary care provider or at facilities that did not match the Veteran’s preferred facility, as these visits may have been for urgent care or other non-longitudinal care.

### Treatment Strategies and Assignment

We compared enrollment in HBPM (intervention) versus usual care (control). HBPM was defined as a monitoring telehealth encounter with an associated diagnosis of hypertension. We defined the start of follow-up as the first eligible primary care visit (i.e., “index visit”, a visit with a systolic BP >140 mmHg with either a systolic BP >140 mmHg at the prior primary care or an average systolic BP >140 mmHg over the previous year). Encounters for telehealth were identified using VHA administrative codes.

### Outcomes and Follow-up

The primary outcomes were all-cause mortality and major adverse cardiovascular events (MACE). Outcomes were assessed from the index date through December 2023. Death was ascertained from the VA vital status file.^16^ Major adverse cardiovascular events, extracted from VINCI, were comprised by emergency department visits or hospitalizations for non-fatal myocardial infarction, stroke, unstable angina, or acute congestive heart failure during follow up. Emergency department visits and hospitalizations for these conditions were identified using diagnosis codes.

### Covariates

Demographic data were accessed from the Patient table in VINCI. Comorbid conditions and illness severity were defined using diagnosis codes and VHA metrics ascertained before the index visit. One-year mortality risk was defined using the validated Care Assessment Need (CAN).^17^ Rurality was determined via the Veterans Affairs urban-rural classification, while socioeconomic status was assessed using the CDC’s Social Vulnerability Index (SVI), mapped at the census tract level through geocoded VINCI data.^18^ Medication use was defined using prescription fill data in VINCI.

### Statistical Analysis

To emulate randomization, individuals in the HBPM group were matched with controls using a weighted greedy matching algorithm with prespecified maximum allowable difference between cases and controls. The variables used in the matching algorithm were ascertained at index date and included age, sex, systolic BP, index encounter date, site of care, CAN score, number of prescribed antihypertensive agent classes, duration of hypertension, and a quarterly specific sufficient dimension reduction summary z score derived from the estimated log odds from a logistic regression of case versus control status. The predictors for the logistic regression for case versus control status included age, gender, race/ethnicity, rurality, index systolic BP, average systolic BP over the prior year, number of prescribed medication classes, most recent serum creatinine level (< 2 mg/dL vs ≥ 2 mg/dL), SVI value, frailty category, CAN score, duration of hypertension, duration of cardiovascular disease, complexity score of the Veteran’s preferred facility, number of patients with hypertension under the care of the Veteran’s primary care provider, Veteran’s primary care provider’s ratio of patients with controlled hypertension, and the Veteran’s primary care provider’s referral rate for hypertension.

Stratified Cox proportional hazards regression models were used, with matched pairs defining the strata, to estimate hazard ratios (HRs) and corresponding 95% confidence intervals (CIs) for the association between HBPM and time-to-event outcomes.^19^ Separate models were fitted for time to MACE and time to mortality. For the MACE analysis, observations were censored at the earliest of death, the study end date (December 31, 2023), or the estimated last observable date within the VHA data. For the mortality analysis, observations were censored at the earlier of the study end date or the estimated last observable date. The estimated last observable date was defined as 25 months after the later of the last recorded measurement date or last recorded visit date.

A stratified Fine–Gray competing-risks model was used for MACE, with death treated as a competing event.^20^ The primary stratified Cox models for MACE and mortality were also adjusted for case management status. Effect modification by case management was evaluated by comparing nested stratified Cox models with and without an HBPM enrollment-by-case management interaction term using a likelihood ratio test.

Subgroup analyses were performed by repeating the stratified Cox regression models within categories of history of cardiovascular disease, sex, age group, race, baseline systolic BP, and CAN score. Subgroup-specific HRs and 95% CIs were estimated for both MACE and mortality. Additional stratified Cox models were fitted using alternative MACE definitions restricted separately to myocardial infarction/stroke, heart failure, and unstable angina.

To evaluate for potential unmeasured confounding, we conducted a secondary analysis using negative control outcomes and empirical calibration.^21^ We selected negative control outcomes that have no biologically plausible relation to BP or HBPM but would be subject to the same differential ascertainment as the primary outcomes: dermatitis, cerumen, appendicitis, and cataract. The negative control outcomes were analyzed using the same stratified Cox models as the primary outcomes. Their log hazard ratios and standard errors were used to estimate an empirical null distribution, which was then applied to obtain calibrated hazard ratios and 95% confidence intervals for MACE and mortality. The calibration analyses were extended to the prespecified subgroup analyses. Analyses were conducted using R 4.4.2.

## Results

### Participant Characteristics

We identified 2,229,120 Veterans with a BP >140 mmHg at a primary care visit occurring between January 1, 2013 and December 31, 2019. After applying full eligibility criteria, the final cohort consisted of 30,203 Veterans enrolled in HBPM (intervention) for which we identified 30,096 non-enrolled controls. The HBPM group had a mean age of 66.8 years, a mean systolic BP of 159.7 mmHg and diastolic BP of 85.8 mmHg and were taking an average of 2.3 antihypertensive medications at enrollment. The standardized mean differences for distributions of all covariates between the HBPM and control groups were less than 0.100 (Table 1).

**Table 1.**
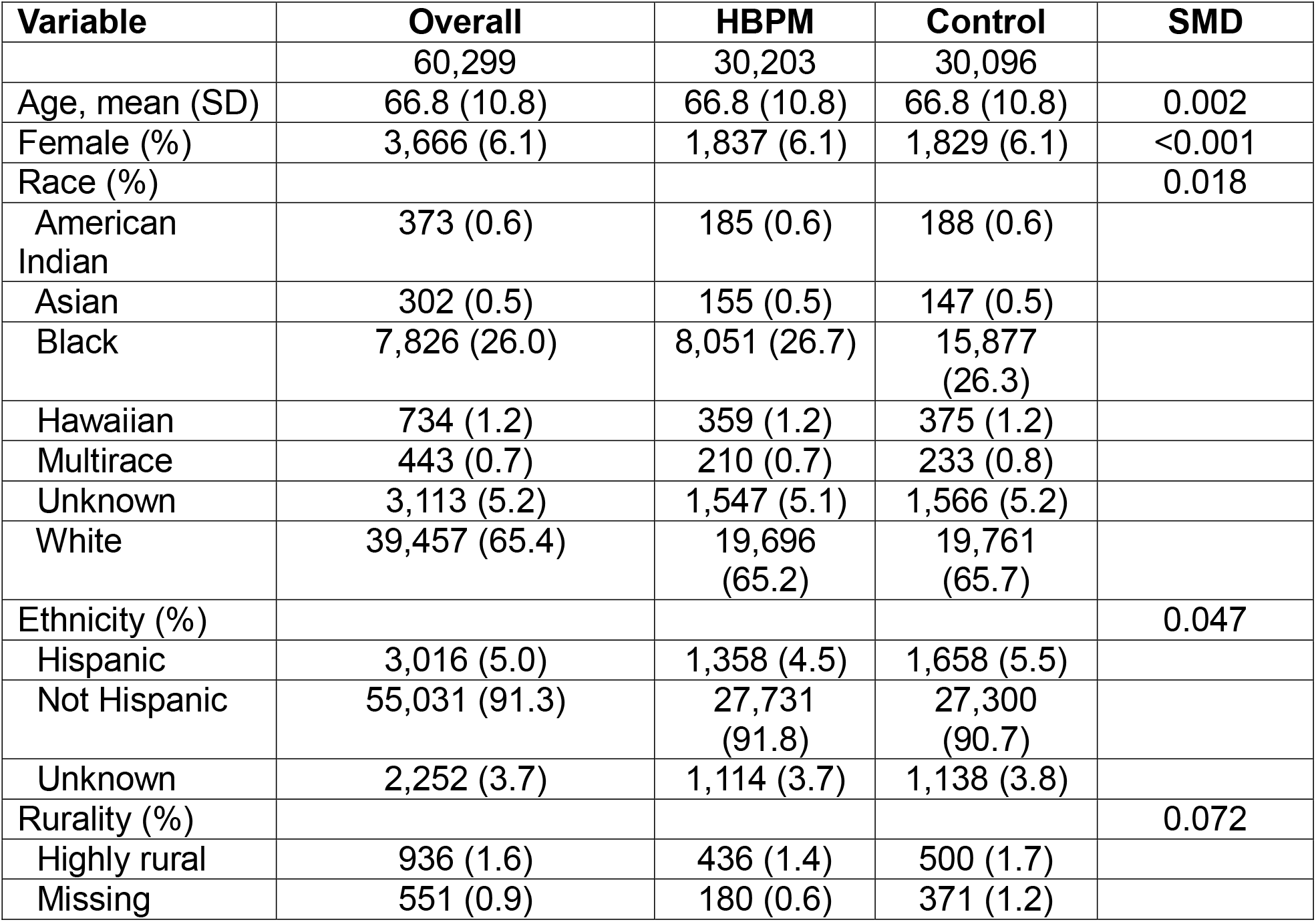
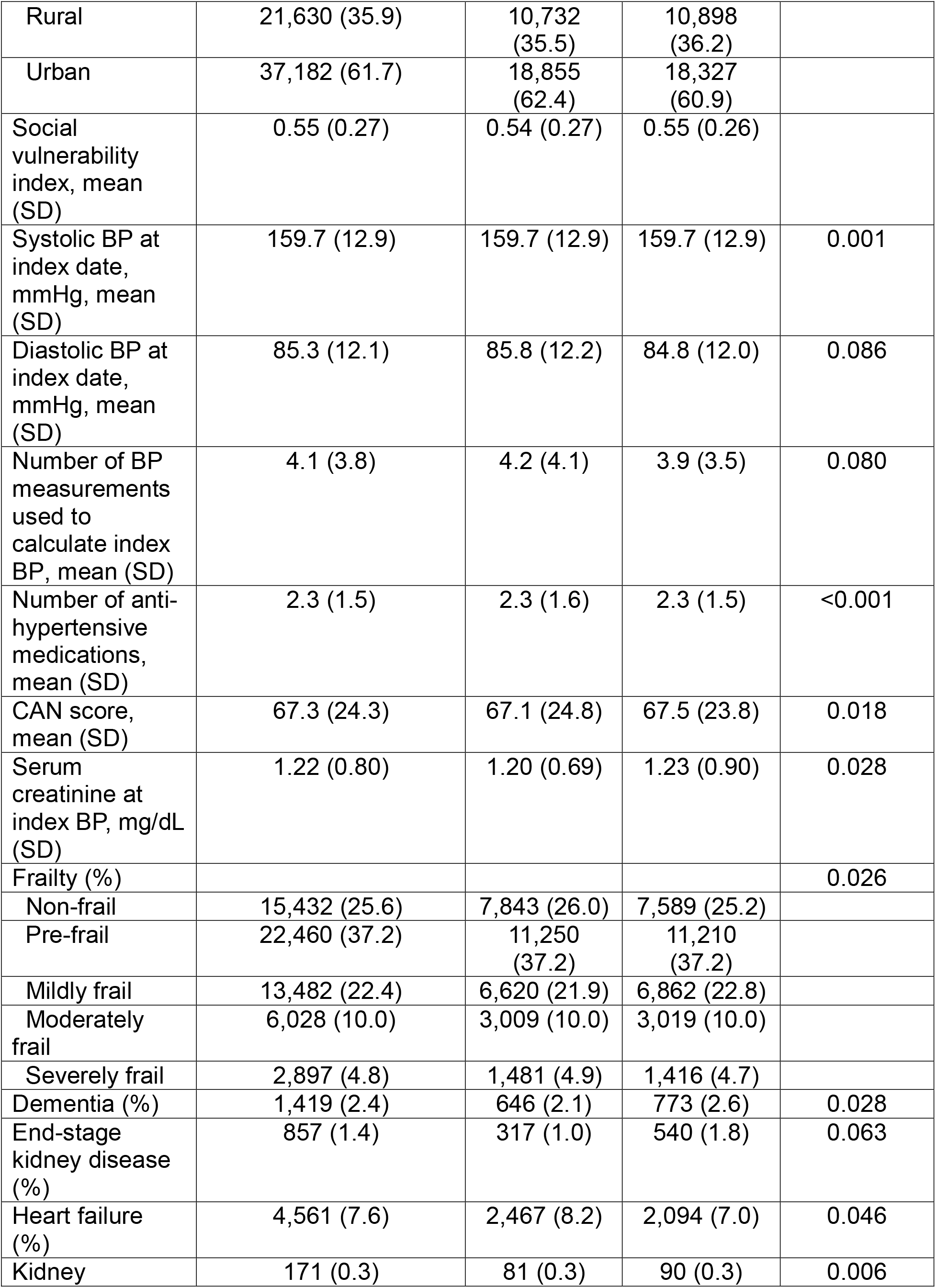
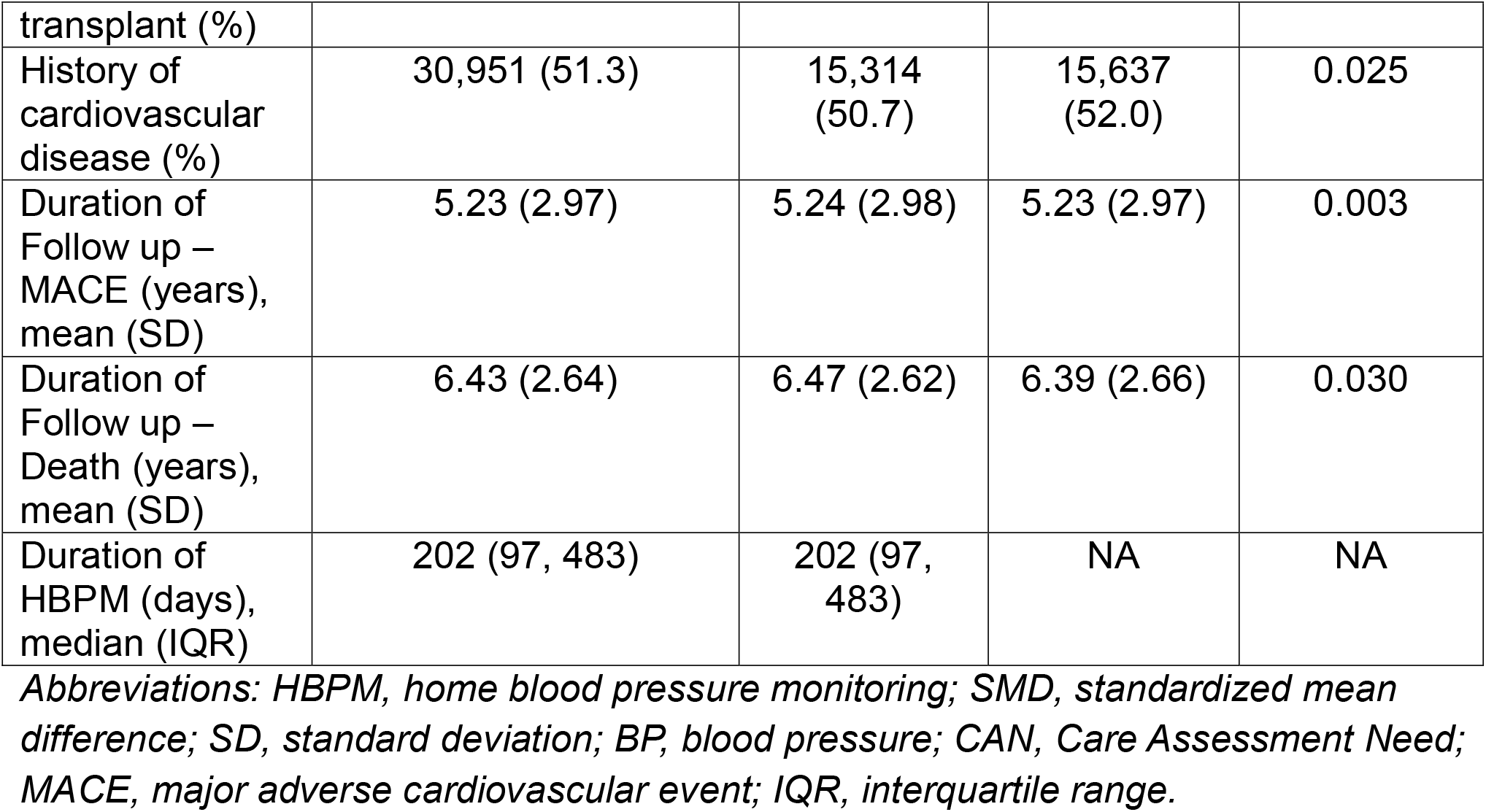
Baseline characteristics of HBPM group and controls.

### Major adverse cardiac events (MACE) and all-cause mortality

MACE occurred in 10,471 (34.7%) individuals in the HBPM group and 10,125 (33.6%) in the control group. Of the MACE events in the HBPM group, 1,838 (17.6%) occurred during the HBPM period and 6,894 (65.8%) occurred after HBPM. HBPM was associated with a 4.1% higher risk of MACE (hazard ratio (HR) 1.04; 95% confidence interval (CI) 1.01, 1.08) (Table 2). This finding was also consistent in an analysis treating death as a competing risk (HR 1.10; 95% CI 1.07, 1.12). In secondary analyses, HBPM was associated with increased risk for negative control outcomes of appendicitis (HR 1.25; 95% CI 0.97, 1.61), cataracts (HR 1.06; 95% CI 1.01, 1.11), dermatitis (HR 1.02; 95% CI 0.95, 1.09), and impacted cerumen (HR 1.08; 95% CI 1.03, 1.13). In analyses calibrated for negative control outcomes, HBPM was no longer associated with increased risk for MACE, myocardial infarction/stroke, heart failure, or unstable angina (Figure 2).

**Table 2.** Results of primary analysis.

| **Outcome** | **Hazard Ratio** | **95% confidence interval** |
| --- | --- | --- |
| MACE | 1.05 | 1.01 – 1.08 |
| MACE (with death as competing risk) | 1.10 | 1.07 – 1.12 |
| Death | 0.92 | 0.89 – 0.95 |

**Figure 1.**
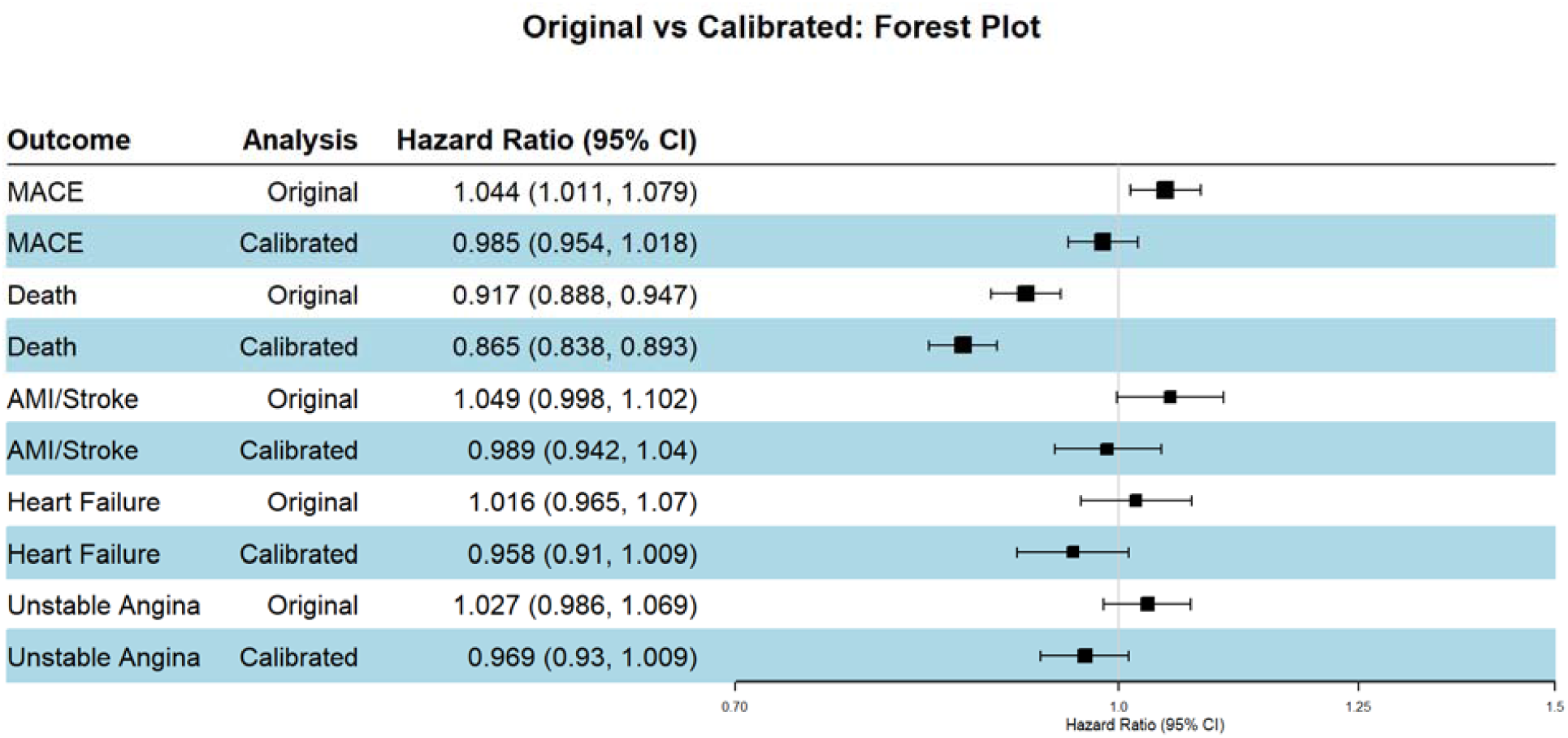
Association between HBPM and outcomes in primary (original) analyses and analyses calibrated for negative control outcomes (calibrated).

**Figure 2.**
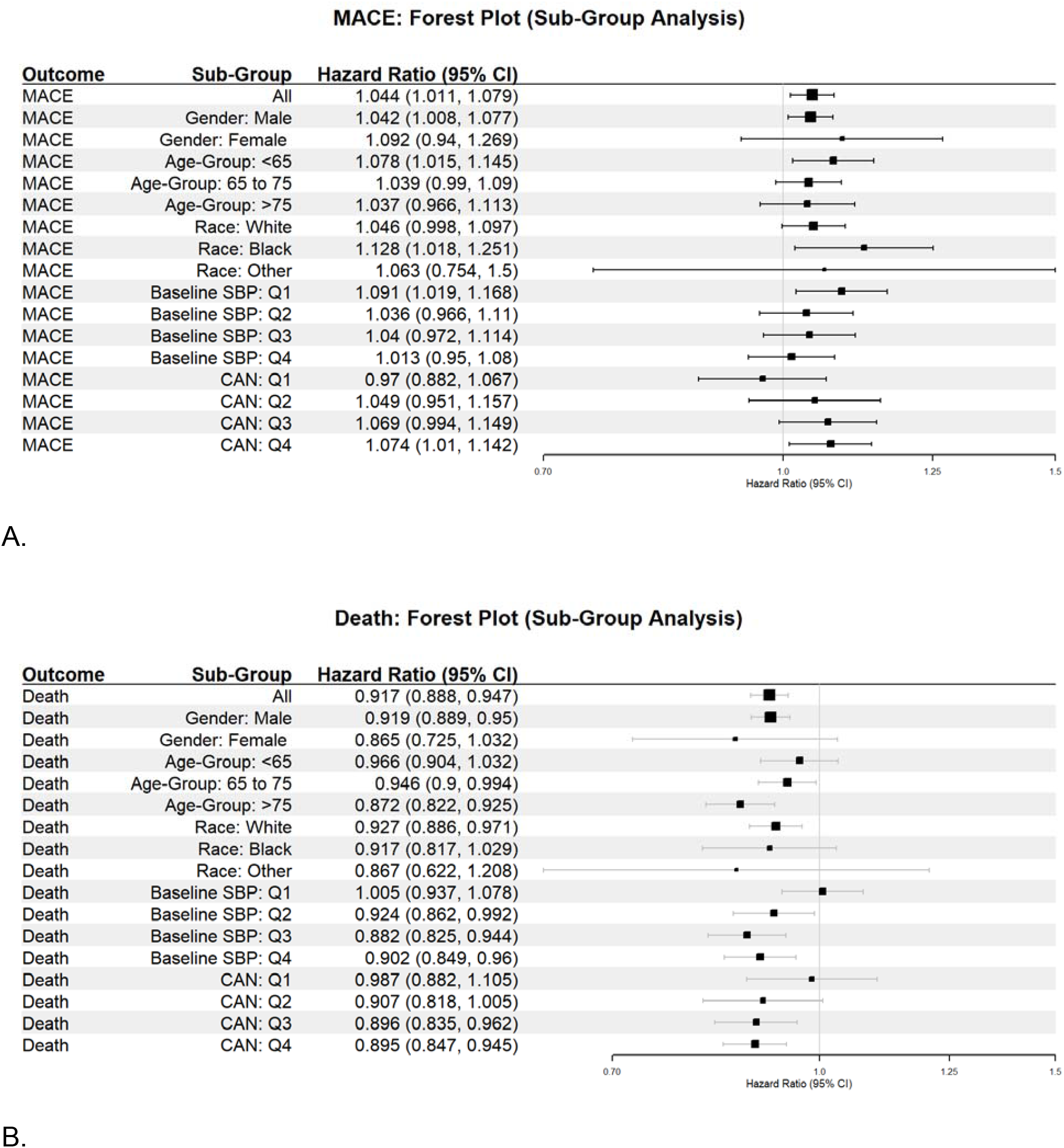
Association between HBPM and MACE (A) and death (B) outcomes among gender, age, race, baseline SBP, and CAN score subgroups.

There were 9,731 deaths (32.2%) in the HBPM group and 10,178 deaths (33.8%) in the control group. HBPM was associated with an 8.3% lower risk of mortality (HR 0.92; 95% CI 0.89, 0.95). In secondary analyses calibrated for negative control outcomes, the magnitude of the reduced risk for mortality was even greater (Figure 1).

### Subgroup analyses: MACE

In subgroup analyses by sex, age group, race, baseline systolic BP, and CAN score, there was no statistically significant difference in the association between HBPM and MACE. However, there was a trend towards increased risk for MACE with lower baseline systolic BP and higher CAN scores (Figure 2A). The increased risk for MACE was present in those with a history of cardiovascular disease (HR 1.06; 95% CI 1.01, 1.11) but not in those without history of cardiovascular disease (HR 0.97; 95% CI 0.90, 1.04). The increased risk for MACE was driven by increased risk for myocardial infarction and stroke (HR 1.05; 95% CI 1.00, 1.10) with a non-significant increased risk for heart failure (HR 1.02; 95% CI 0.97, 1.07) and unstable angina (HR 1.03; 0.99, 1.07).

### Subgroup analyses: Mortality

In subgroup analyses by sex, age group, race, baseline systolic BP, and CAN score, the association between HBPM and lower all-cause mortality was generally consistent, except among those with the lowest baseline systolic BP (HR 1.00 (95% CI 0.94, 1.08) and those with the lowest baseline mortality risk as defined by CAN score (HR 1.00 (95% CI 0.89, 1.12)(Figure 2B).

## Discussion

In this national target trial emulation study of US Veterans with uncontrolled BP, enrollment in HBPM was associated with lower all-cause mortality and higher risk of major adverse cardiovascular events, though the latter was not seen in calibrated analyses. To our knowledge, this is the first study to examine the impact of HBPM in routine clinical care on adverse outcomes.

Meta-analysis of clinical trials have shown that HBPM lowers BP compared to usual care.^12^ Lowering BP in individuals with uncontrolled hypertension reduces their risks of cardiovascular events and mortality.^6–8^ The reduction in mortality we observed in this study demonstrates the effect of HBPM in routine care, providing further support for the 2025 AHA/ACC BP management guideline on use of out-of-office BP to manage hypertension.^10^ Additionally, our findings highlight the potential public health impact with increasing utilization of HBPM, as even modest reductions in mortality translate to large absolute numbers of lives when applied to a condition as prevalent as hypertension.

Our study’s finding that HBPM was associated with increased MACE risk was unexpected. There are a number of possible contributing factors to this finding. First, it may be that HBPM reduced the severity of MACE events, and events that would otherwise have been fatal would thus be counted in the MACE outcome (which was only non-fatal CV events) while lowering mortality via decreased fatal CV events.

However, we also suspect that differential ascertainment likely contributed to the association between HBPM and increased risk of MACE, a theory which is supported by the negative control outcome analyses. We hypothesize that patients enrolled in HBPM may have had more frequent contact with the medical system, potentially leading to more opportunities for outcome detection. After adjusting for the negative control outcomes in the calibrated analysis, there was no longer a significant association between HBPM and MACE. Additionally, other residual confounding may have also contributed to the observed increase in MACE risk. Subgroup analyses showed that the increased risk of MACE was driven by those with those with a known history of cardiovascular disease prior to enrollment. It is possible that clinicians providing HBPM may have had a lower threshold to recommend emergency care to patients with known cardiac history. Finally, although the distribution of baseline heart failure was well-balanced between patients enrolled in HBPM and the control group, we used administrative codes to define baseline heart failure, which do not include heart failure severity (i.e., New York Heart Association class). Thus, some patients in the enrolled group may have had more severe or “active” heart failure and their clinicians may have enrolled them for monitoring of both blood pressure and heart failure.

The current study has several strengths. The target trial emulation study design is a strength of our study, as is outcome ascertainment through the VHA system and linked Medicare data, which minimizes loss to follow up. Limitations of this study include the low proportion of females, reliance on administrative codes to define cardiovascular outcomes, which are highly specific but may lack sensitivity, and healthy user bias among those who enrolled in HBPM, though the latter seems unlikely given the high baseline cardiovascular disease risk in this cohort. Additionally, not all patients who enrolled in HBPM actually had home BP measured. We were also unable to identify team-based care, which influences BP control in patients enrolled in HBPM.

In summary, in this large target trial emulation study of United States adults with uncontrolled hypertension, HBPM was associated with lower all-cause mortality. After accounting for differential outcome ascertainment, there was no significant association with MACE. These findings support wider implementation of HBPM in routine care and warrant further study of the mechanisms through which HBPM reduces mortality.

## Data Availability

The data used in this study are clinical datasets that cannot be deidentified and are property of the United States Government and the Department of Veterans Affairs, thus these data may not be shared outside of the VA.

## Notes

**Conflict of Interest Statement:** The authors report no relevant conflicts of interest to disclose.

### Competing Interest Statement

The authors have declared no competing interest.

### Author Declarations

The Institutional Review Board of University of Minnesota gave ethical approval for this work (approval number STUDY00014620). The Institutional Review Board of the Minneapolis Veterans Health Administration gave ethical approval for this work (approval number 4739-B).

### Funding

- National Heart Lung and Blood Institute: NHLBI 5R01HL157667

